# Challenges of false positive and negative results in cervical cancer screening

**DOI:** 10.1101/2020.03.17.20037440

**Authors:** David Robert Grimes, Edward M.A. Corry, Talía Malagón, Ciaran O’Riain, Eduardo L. Franco, Donal J. Brennan

## Abstract

**Objective:** To quantify the impact and accuracy of different screening approaches for cervical cancer, including liquid based cytology (LBC), molecular testing for human papillomavirus (HPV) infection, and their combinations via parallel co-testing and sequential triage. The secondary goal was to predict the effect of differing coverage rates of HPV vaccination on the performance of screening tests and in the interpretation of their results.

**Design:** Modelling study.

**Main outcomes measured:** Different screening modalities were compared in terms of number of cases of Cervical intra-epithelial neoplasia (CIN) grade 2 and 3 detected and missed, as well as the number of false positives leading to excess colposcopy, and number of tests required to achieve a given level of accuracy. The positive predictive value (PPV) and negative predictive value (NPV) of different modalities were simulated under varying levels of HPV vaccination.

**Results:** The model predicted that in a typical population, primary LBC screening misses 4.9 (95% Confidence Interval (CI) 3.5-CIN 2 / 3 cases per 1000 women, and results in 95 (95% CI: 93-97%) false positives leading to excess colposcopy. For primary HPV testing, 2.0 (95% CI: 1.9-2.1) cases were missed per 1000 women, with 99 (95% CI: 98-101) excess colposcopies undertaken. Co-testing markedly reduced missed cases to 0.5 (95% CI: 0.3-0.7) per 1000 women, but at the cost of dramatically increasing excess colposcopy referral to 184 per 1000 women (95% CI: 182-188). Conversely, triage testing with reflex screening substantially reduced excess colposcopy to 9.6 cases per 1000 women (95% CI: 9.3 - 10) but at the cost of missing more cases (6.4 per 1000 women, 95% CI: 5.1 - 8.0). Over a life-time of screening, women who always attend annual and 3-year co-testing were predicted to have a virtually 100% chance of falsely detecting a CIN 2 / 3 case, while 5 year co-testing has a 93.8% chance of a false positive over screening life-time. For annual, 3 year, and 5 year triage testing (either LBC with HPV reflex or vice versa), lifetime risk of a false positive is 35.1%, 13.4%, and 8.3% respectively. HPV vaccination rates adversely impact the PPV, while increasing the NPV of various screening modalities. Results of this work indicate that as HPV vaccination rates increase, HPV based screening approaches result in fewer unnecessary colposcopies than LBC approaches.

**Conclusion:** The clinical relevance of cervical cancer screening is crucially dependent upon the prevalence of cervical dysplasia and/or HPV infection or vaccination in a given population, as well as the sensitivity and specificity of various modalities. Although screening is life-saving, false negatives and positives will occur, and over-testing may cause significant harm, including potential over-treatment.

## Introduction

The impact of cervical cancer screening programmes with conventional cytology has been dramatic, as shown in numerous national programmes. While its effect has been primarily restricted to squamous cell carcinoma (SCC) in women over the age of 25, the estimated 80% reduction in mortality in high-quality national screening programs illustrates its truly positive impact(1–3). This ability to save women’s lives has been key in justifying the substantial cost associated with screening, which in 2004 was estimated at £36,000 per life saved in the UK(1).

The evolution of cervical cancer screening to include HPV testing is a desirable step (4). However, this evolution necessitates that critical lessons from history are not repeated. HPV DNA testing as a screening tool has already demonstrated superior sensitivity in detecting cervical intra-epithelial neoplasia (CIN) grade 2 and 3(5). Less emphasis has been placed on the high prevalence of HPV infection, and that sub-optimal implementation of primary HPV screening will result in increased referrals to colposcopy. This is especially relevant as national programmes transition to HPV testing, which necessitates re-education of the screened population regarding the benefits and limitations of cervical screening, particularly with the meaning of a “normal smear” (6).

A central consideration is the disproportionate impact false negatives can have on women who attend for cervical screening and the need to determine what represents acceptable false negative rates within programmes (7,8). This is further complicated by the legal standard in the UK and Ireland that screeners have “absolute confidence” in a negative test (9,10).

Screening tests are however far from perfect in the identification of disease. Cochrane review figures (11) indicate that for every 1000 women screened 20 will be diagnosed with cervical intra-epithelial neoplasia (CIN) of grades 2 or 3 (CIN2/3). Cytology alone will identify 15 women of these women and screening with HPV testing would identify an additional 3 that would have been missed with cytology (18 vs 15). This increased sensitivity of HPV testing comes at the expense of lower specificity than liquid-based cytology (LBC). Typically, HPV screening results in an additional 4 women in 1000 being told they have a suspected lesion with the subsequent potential for unnecessary, harmful interventions.

It is also worth looking at the future of screening and at the influences that will affect its performance. HPV vaccination is already markedly reducing the prevalence of CIN across the world. As uptake increases, prevalence of HPV infection drops. While relatively recent, HPV vaccination is having a dramatic impact; a recent study in a Scottish cohort found a reduction of 88% in cervical disease due to vaccination (12), and modelling studies suggest Australia could virtually eliminate cervical cancer in coming decades due to its pioneering adoption and long-term maintenance of a successful vaccination program (13). As prevalence falls, a positive result is increasingly likely to not reflect a true case. The positive predictive value (PPV) of any test is the probability that a positive result truly reflects underlying disease that needs to be discovered and treated. There is already evidence of vaccination in younger women decreasing PPV (12,14), which was predicted at the time the HPV vaccine was first approved (15). Theoretically, one would expect an impact on negative predictive value (NPV), the likelihood that a negative result can be safely reassuring that no disease is present. Implications of increased vaccination uptake must also be considered for any viable screening modality, as these shape interpretation of results and clinical judgement.

In this work, we use a mathematical model to examine the nuances of screening programmes using different screening modalities, examining the impact of different strategies on CIN2/3 cases detected, missed, and excess referral to colposcopy. We demonstrate the impacts of strategies to reduce false negative rates and examine whether these approaches might potentially result in overtreatment. Finally, we also examine the impact of HPV vaccination on screening accuracy, and likely challenges for the future to maintain beneficial screening programs.

## Methods

### Screening considerations

Screening is performed on asymptomatic women, in the hope of detecting potential CIN2/3 lesions before they advance into cancer. There are three especially relevant parameters to any screening test and in our model: prevalence, sensitivity and specificity.

- Prevalence (p) - The fraction of a given population that have a disease (in this case, CIN 2/3)
- Sensitivity (s*n*) - The proportion of cases with disease (CIN2+ in this case) that are correctly identified as positive by the screen. A test with a high sensitivity has in consequence a low false negative rate.
- Specificity (s*p*) - The proportion of individuals without disease correctly identified as such by screening, i.e., negative. A test with a high specificity means a test has a low rate of false positives.

Tests do not have perfect sensitivity nor specificity. Consequently, false positive results (incorrectly identifying lesion-free women as CIN2/3 or worse) and false negative results (missing instances of CIN2/3 or cancer) are unavoidable in practice. Their impact on clinical decisions is a function of sensitivity and specificity of the test, and prevalence of disease itself. The PPV and NPV are respectively the probability that a test positive is a true positive and that a test negative is a true negative and are defined in more detail in the mathematical appendix. As prevalence falls, a positive result is increasingly likely to not reflect a true case, and PPV would thus decline. Conversely, as the prevalence of a disease decreases, the NPV of testing tends to increase.

### Testing modalities and implementation

#### Primary testing, triage, and co-testing

Worldwide, there are numerous different approaches to cervical screening, and these can differ even inside a country. We will thus concentrate on general methods for illustration. LBC Primary test-only is illustrated in figure 1(a), where a positive LBC test is referred to colposcopy. Conversely, primary LBC may be performed with reflex HPV test, where a ASC-US positive results on a smear are then HPV tested. For primary HPV testing, a positive result is followed by cytological examination of the same specimen, where If abnormalities are then detected, referral is made to colposcopy. Another option with some clinical use is co-testing, where both HPV and LBC tests are performed. Worldwide, there are different ways to manage co-test results, but only interpretation is that a positive result in either arm instigates an elevation to colposcopy, illustrated in figure 1(b). Finally, reflex triage approaches are illustrated in figure 1(c). where an LBC screen can be performed, and AS-CUS results interrogated with a reflex HPV test, with positive results referred to colposcopy, as is common in Australia. Alternatively, the converse can occur (HPV test with reflex LBC, as is the recommended case in Ireland). In this work, we simulate all these general approaches.

**Figure 1.**
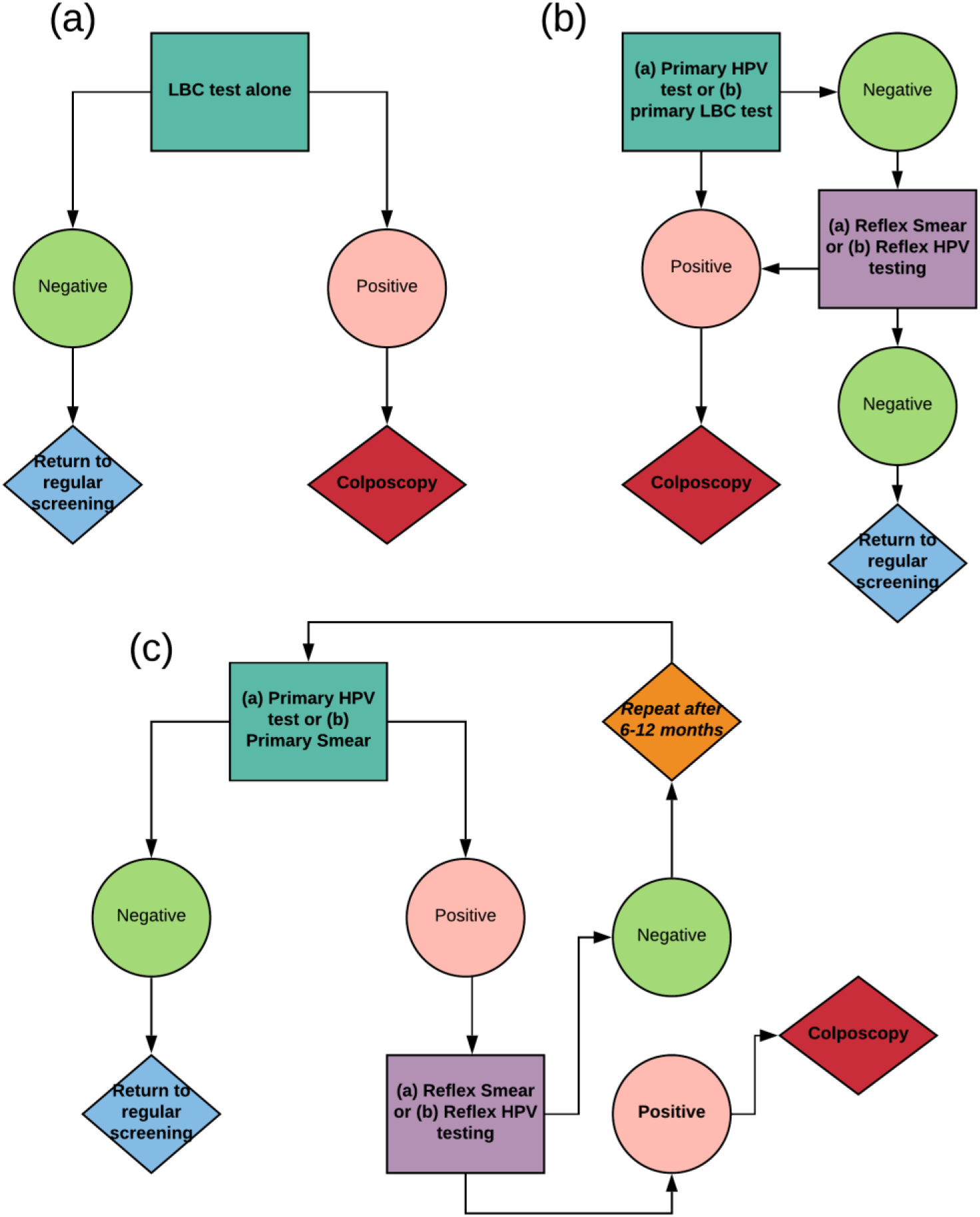
Flowcharts for (a) Primary testing only (b) Co-testing (c) Triage testing. Note that these are general schemas, and that there is large variation worldwide in exact implementation.

### Repeat testing of negative results

Another potential option to reduce the risk of false negatives is to perform n retests of tests that are initially negative. As detailed in the mathematical appendix, multiple testing with LBC modalities constantly reduces the number of missed cases of CIN2/3 cells, but at the cost of increasing excess unnecessary colposcopy (increased false positives). To the author’s knowledge such a methodology hasn’t been previously implemented in precisely such a fashion, but it is worth noting that repeat testing of negative results is similar to asking women who screen negative to come back for another regular screen after a certain interval, similar to what is recommended in most countries. While the model here does not attempt to reproduce the natural history of HPV infection, it can still be employed to illuminate the clinical value and untoward consequences of multiple screenings for a woman over time if she has cervical disease or not.

### The impact of the HPV vaccine on screening accuracy

The HPV vaccine will markedly reduced CIN prevalence. A reduction in prevalence, however, has implications for both the PPV and NPV of screening tests. To model the impact of vaccine uptake on screening performance, we took predictive data from 29 modelling studies as previously described in the literature(16) to estimate the expected HPV prevalence under different levels of vaccine uptake (presuming vaccination against subtypes 6,11, 16 and 18), and then calculated PPV and NPV for both HPV testing and LBC.

### Brief Modelling description

A mathematical model was constructed to simulate likely outcomes of different modalities and implementations, so that they could be cross-compared. Full mathematical details of the model are in the appendix, as well as details on parameter estimation. Outcomes were simulated for a hypothetical cohort of 1000 women with a CIN2+ prevalence of 2%. This is a simplification, as natural history models show significant variation in CIN2+ prevalence with age and nationality(17), but this figure for prevalence is broadly representative and appropriate to assess screening performance in a randomly selected cohort(11). Table 1 lists the parameter values used for simulations. In addition to test outcomes, the model is also capable of yielding cumulative life-time probability of a false positive for women without CIN 2/3 as a function of test and testing interval, for the scenario where her underlying health status does not change. Exact formula are given in the supplementary material, and using these methods, we also model the impact of different screening regimens in a typical population to compare their false negative and false positive risk as a function of screening frequency.

**Table 1.** Prevalence, sensitivity, and specificity of CIN2/3 tests

| Parameters | Estimate with 95% CI where available |
| --- | --- |
| Prevalence of CIN2/3 in typical population(11) | 2.0 (-) |
| Prevalence of detectable HR-HPV in typical population <sup>a</sup> | 8.4 (-) |
| CIN2/3 attributable to testable hr-HPV | 95.0 (-) |
| CIN2/3 Sensitivity (LBC)(11) | 75.5 (66.6 - 82.7) |
| CIN2/3 Specificity (LBC)(11) | 90.3 (90.1-90.5) |
| CIN2/3 Sensitivity (HPV testing)(11) | 89.9 (88.6-91.1) |
| CIN2/3 Specificity (HPV testing)(11) | 89.9 (89.7-90.0) |
| HR-HPV test Sensitivity(18) | 94.7 (-) |
| HR-HPV test Specificity <sup>b</sup> | 96.0 (95.7-96.1) |

<sup>a</sup> Estimated value(18–21). See appendix for details.
<sup>b</sup> Derived value. See appendix for details. CIN: cervical intraepithelial neoplasia; HR-HPV: high-risk human papillomaviruses; LBC: liquid-based cytology;

### Model outcomes

Different implementations were tested, with emphasis on the following aspects vital to any screening programme

1. Lesions missed per 1000 women (false negatives).
2. Theoretical maximum excess colposcopy referrals per 1000 women screened (false positives leading to unnecessary colposcopy referral)
3. The positive predictive value of a given implementation.
4. The negative predictive value of a given implementation.
5. Total number of tests undertaken per 1000 women.

Ideally, false negative (FN) rates should be as low as reasonably possible, so that the NPV can approach 100%. In practice this is impossible (see mathematical supplementary material), but in principle one could reduce the number of FNs by retesting negative results. We might instead define an appropriate threshold for “absolute” confidence in a negative result, below which we can conclude with extremely high certainty that a woman does not have CIN2+. As an example in our study, we selected a confidence threshold of *t*_*s*_ < 0.1%, which is consistent with an NPV of greater than 99.99%, and is the threshold likely to be the foundation of some forthcoming US guidelines(22). Each screening round, however, CIN2/3 prevalence in the cohort changes from po initially to *p*(*n*). This acts to reduce missed cases of CIN 2/3, but conversely increases false positives requiring colposcopy. The formula for this is outlined in the mathematical appendix. Here, we simulate likely impacts of cautionary re-testing of negatives, ascertaining how many screening rounds would be required to achieve a threshold of *t*_*s*_ and the implications for colposcopy rates.

## Results

### Cross comparison of screening modalities and implementations

Table 2 shows the modelled comparison of screening statistics for different modalities discussed. Due to low prevalence of CIN2/3, NPV is consistently high across all modalities (> 99% typically) with the exception of screening modalities with triage testing, where it is lower. Triage testing leads to more missed CIN2/3 cases, but drastically reduces excess colposcopy referrals; HPV with LBC reflex triage results in typically only 9.6 excess colposcopy referrals per 1000 women screened, but increases the number of false negatives (6.4 per 1000 women screened). For primary testing alone as illustrated in figure 2, HPV testing detects 19% more cases than LBC testing, but led to 4% more colposcopy referrals relative to LBC.

**Table 2.**
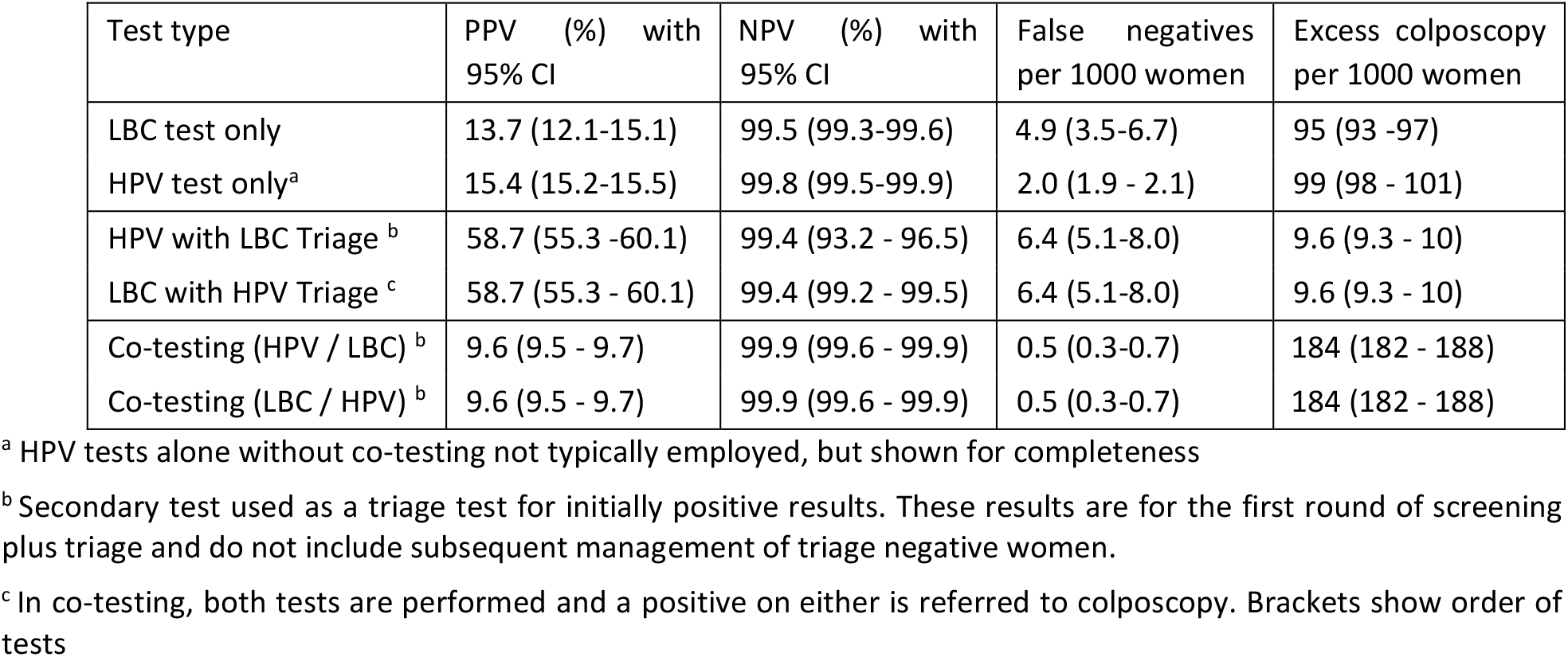
CIN2/3 detection statistics for cohort of 1000 women

| Test type | PPV (%) with 95% CI | NPV (%) with 95% CI | False negatives per 1000 women | Excess colposcopy per 1000 women |
| --- | --- | --- | --- | --- |
| LBC test only | 13.7 (12.1-15.1) | 99.5 (99.3-99.6) | 4.9 (3.5-6.7) | 95 (93 -97) |
| HPV test only<sup>a</sup> | 15.4 (15.2-15.5) | 99.8 (99.5-99.9) | 2.0 (1.9 - 2.1) | 99 (98 - 101) |
| HPV with LBC Triage<sup>b</sup> | 58.7 (55.3 -60.1) | 99.4 (93.2 - 96.5) | 6.4 (5.1-8.0) | 9.6 (9.3 - 10) |
| LBC with HPV Triage<sup>c</sup> | 58.7 (55.3 - 60.1) | 99.4 (99.2 - 99.5) | 6.4 (5.1-8.0) | 9.6 (9.3 - 10) |
| Co-testing (HPV / LBC)<sup>b</sup> | 9.6 (9.5 - 9.7) | 99.9 (99.6 - 99.9) | 0.5 (0.3-0.7) | 184 (182 - 188) |
| Co-testing (LBC / HPV)<sup>b</sup> | 9.6 (9.5 - 9.7) | 99.9 (99.6 - 99.9) | 0.5 (0.3-0.7) | 184 (182 - 188) |

<sup>a</sup> HPV tests alone without co-testing not typically employed, but shown for completeness
<sup>b</sup> Secondary test used as a triage test for initially positive results. These results are for the first round of screening plus triage and do not include subsequent management of triage negative women.
<sup>c</sup> In co-testing, both tests are performed and a positive on either is referred to colposcopy. Brackets show order of tests

**Figure 2.**
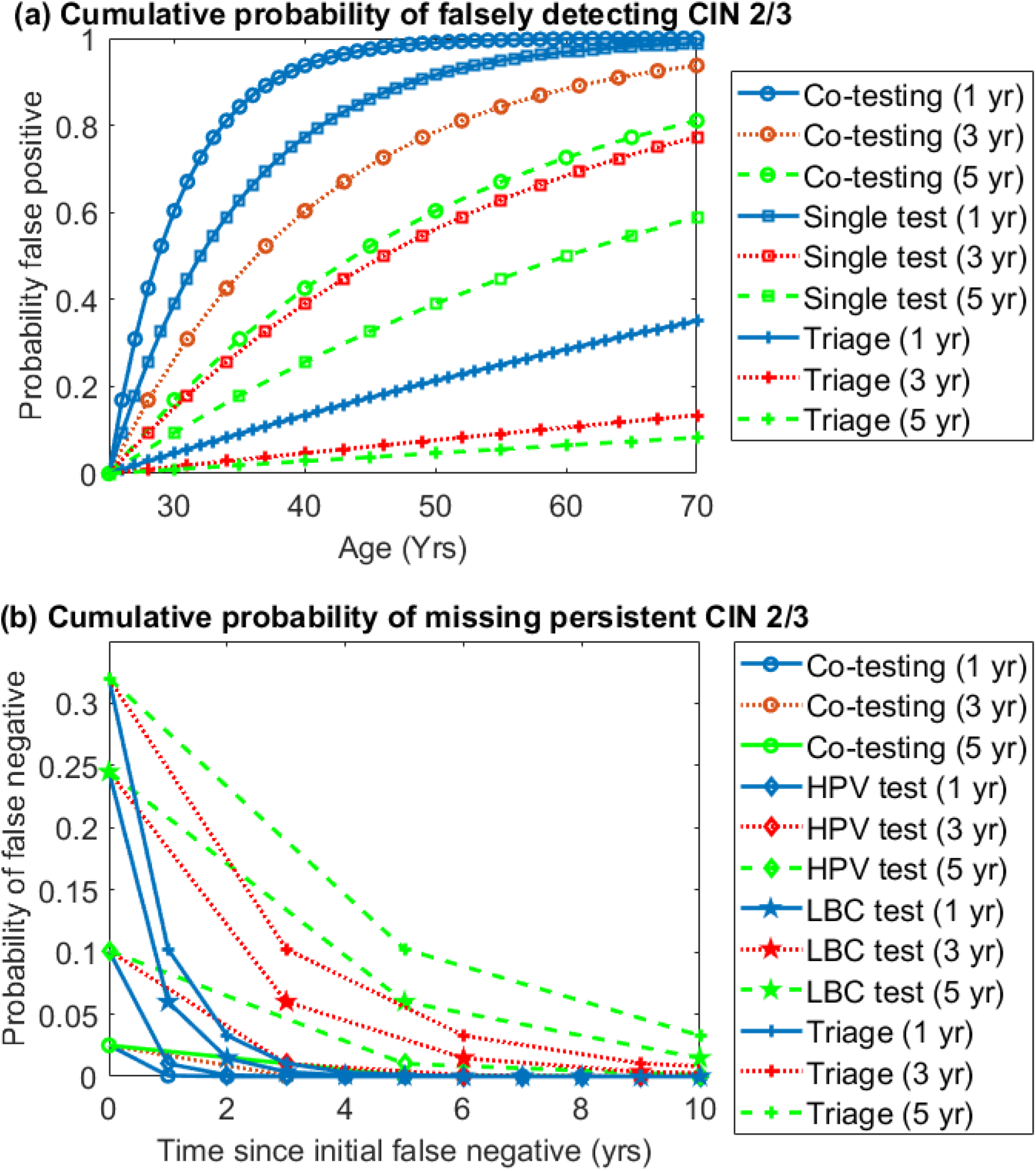
(a) Cumulative probability of getting a false positive result with different modalities and screening intervals over a patient life-time, assuming screening begins at 25 and ends at 70. Note that results for single LBC / HPV tests are so similar they have been collapsed into one category (single test) for clarity, with caveat that HPV-only screening is not typically performed and shown only for completeness. (b) Cumulative probability of missing persistent CIN 2/3 for different modalities with different screening intervals, with the horizontal axis depicting number of years since the initial false negative result. Note that HPV only screening is shown for comparison despite limited clinical use.

Primary testing however produces an abundance of false positives. This can be reduced with triage testing, as depicted in figure 2. Both triage modalities (HPV with LBC reflex or LBC with HPV reflex) lead to only 10% of the false positives of LBC, at the cost of detecting only 90% of the CIN 2/3 cases LBC would detect. More importantly, triage outcomes are the same regardless of the primary test. This is relevant, as it may have economic impact. For example, 1000 women with HPV primary would require 1000 HPV tests, and a follow up of 117 triage LBC tests. Conversely, an LBC primary with HPV reflex for 1000 women requires 1000 LBC and 110 follow up triage HPV tests, as derived from formula in mathematical appendix.

Co-testing can markedly increases the number of cases detected, detecting 29% more cases than LBC alone. This however comes at the cost of increasing the false positive rate by 94%. Table 3 shows the number of test iterations required to reach an NPV of greater than 99.99%. This analysis does not account for test interval or triage strategies, discussed below.

**Table 3.** Screening rounds required to achieve t_s_ < 0.1% (NPV > 99.99%)

| Test type<sup>a</sup> | Screening rounds required | Total tests required per 1000 women<sup>b</sup> | Missed positives per 1000 women | Excess colposcopy per 1000 women |
| --- | --- | --- | --- | --- |
| Liquid-based cytology | 3 | 2690 (2688 - 2694 ) | 0.3 (0.1-0.7) | 258 (254 - 263) |
| HPV test only<sup>c</sup> | 2 | 1883 (1881-1884) | 1 (0.9-1.0) | 138 (136 - 142) |
| Co-testing (HPV / LBC) | 1 (combined) | 1883 (1881 -1884) | 0.5 (0.3-0.7) | 184 (182 - 188) |
| Co-testing (LBC / HPV) | 1 (combined) | 1890 (1890 -1890) | 0.5 (0.3 - 0.7) | 184 (182 - 188) |

<sup>a</sup> Triage results not shown, as repeated triage does not increase NPV. See text for details.
<sup>b</sup> Total tests is the number of tests of initial type (1000) plus follow-up type. For example, Co-testing HPV/LBC implies 1000 HPV tests (initial) plus 883 (881-884) LBC tests.
<sup>c</sup> HPV tests alone without co-testing not typically employed, but shown for completeness

### Implications of test modality and frequency on over-screening and missed positives

The accuracy of screening is also dependent on inter-test interval. Figure 2 shows outcomes for different modalities, depicting both the cumulative probability of a false positive result over a screening lifetime (assuming screening begins at 25 and ceases at 70) for a CIN2/3 negative woman and the probability of missing successive true and persistent CIN 2/3 (Figure 2b). Intervals of 1, 3, and 5 years are shown. As can be seen clearly, triage testing results in much fewer false positives, but at the cost of missing a greater proportion of CIN2/3 cases. Co-testing by contrast, can drastically reduce the number of CIN2/3 cases missed, but at the cost of increasing the false positive rate markedly. This can be somewhat ameliorated by reducing screening frequency.

Triage tests themselves have some added nuance that must be considered. Table 4 depicts the likely outcome of HPV primary testing with LBC reflex, considering the expedited retesting process that results from a positive HPV infection status. In these instances, women are tested again 6-18 months after this result. The retest can take several forms; another HPV test (UK(23) and Australia(24)), an LBC (The Netherlands(25)) or both (United States(26)). Outcomes and times to detection are shown in table 4, which clearly illustrates that retesting with both HPV and LBC detects more cases than a HPV retest alone, and much more cases relative to a single LBC retest. Another important consideration for Triage tests is the primary test; while outcomes are the same, the order of testing slightly impacts the total number of tests undertaken, as shown in table 3. Depending on the cost differential between HPV and LBC screens, this might be economically relevant, as explored further in discussion.

**Table 4.** Possible Triage outcomes with expedited retesting for a woman with CIN 2/3

| Triage type | Probability %<br>(95% CI) | Outcome | Time to CIN2/3 detection |
| --- | --- | --- | --- |
| **Initial Screening (HPV / LBC triage)** |  |  |  |
| HPV and CIN2/3 cytology positive | 71.5 (63.1-78.4) | Colposcopy | Immediate |
| HPV detected, CIN2/3 cytology negative | 23.2 (16.4-31.6) | Expedited retest | Expedited test dependent |
| HPV missed, no cytology triage | 5.3 (-) | False negative | Next screening cycle at earliest |
| **Expedited retest (HPV Only)** |  |  |  |
| HPV detected | 94.7 (-) | Colposcopy | 6-18 months after initial screen |
| HPV missed | 5.3 (-) | False negative | Next screening cycle at earliest |
| *Probability of missing CIN2/3+ after retest* | 1.2 (0.9 - 1.7) |  |  |
| **Expedited retest (LBC Only)** |  |  |  |
| CIN2/3 detected | 75.5 (66.6 - 82.7) | Colposcopy | 6-18 months after initial |
| CIN2/3 missed | 24.5 (17.3 - 33.4) | False negative | Next screening cycle at earliest |
| *Probability of missing CIN2/3+ after retest* | 5.7 (2.8 - 10.6) |  |  |
| **Expedited retest (HPV and LBC)** |  |  |  |
| Either HPV or CIN2/3 detected | 98.7 (98.2 - 99.1) | Colposcopy | 6-18 months after initial |
| Both HPV and CIN2/3 missed | 1.3 (0.9-1.8) | False negative | Next screening cycle at earliest |
| *Probability of missing CIN2/3+ after retest* | 0.3 (0.2 - 0.6) |  |  |

Table 3 and Figure 2 also show that while multiple screening rounds increase detection rate, it does so at the cost of increasing over-diagnosis and expense. For example, three rounds of LBC improves detection by 30% relative to a single screen, but requires a factor of 2.69 more tests, with 172% the false positive rate, rendering it an unsustainable approach.

### Impact of the HPV vaccine on screening accuracy

Figure 3(a) and 3(b) depict the impact of screening on the number of false positives (excess colposcopy) for HPV, LBC, and Triage testing as vaccination rates increase. Figure 3(c) and (d) show the PPV and NPV change with vaccination rates for HPV and LBC tests. Confidence envelopes were derived from a selection of natural history models as previously described. As vaccination rates increase, PPV of both tests fall, while NPV rises. What is immediately clear, however, is that HPV testing is a superior modality as vaccination increases, as it results in much fewer false positives than LBC testing. This can further be improved by implementing triage testing, and the results of this simulation would strongly suggest HPV testing is a superior method of screening to employ as vaccination rates increase and HPV infection decreases.

**Figure 3.**
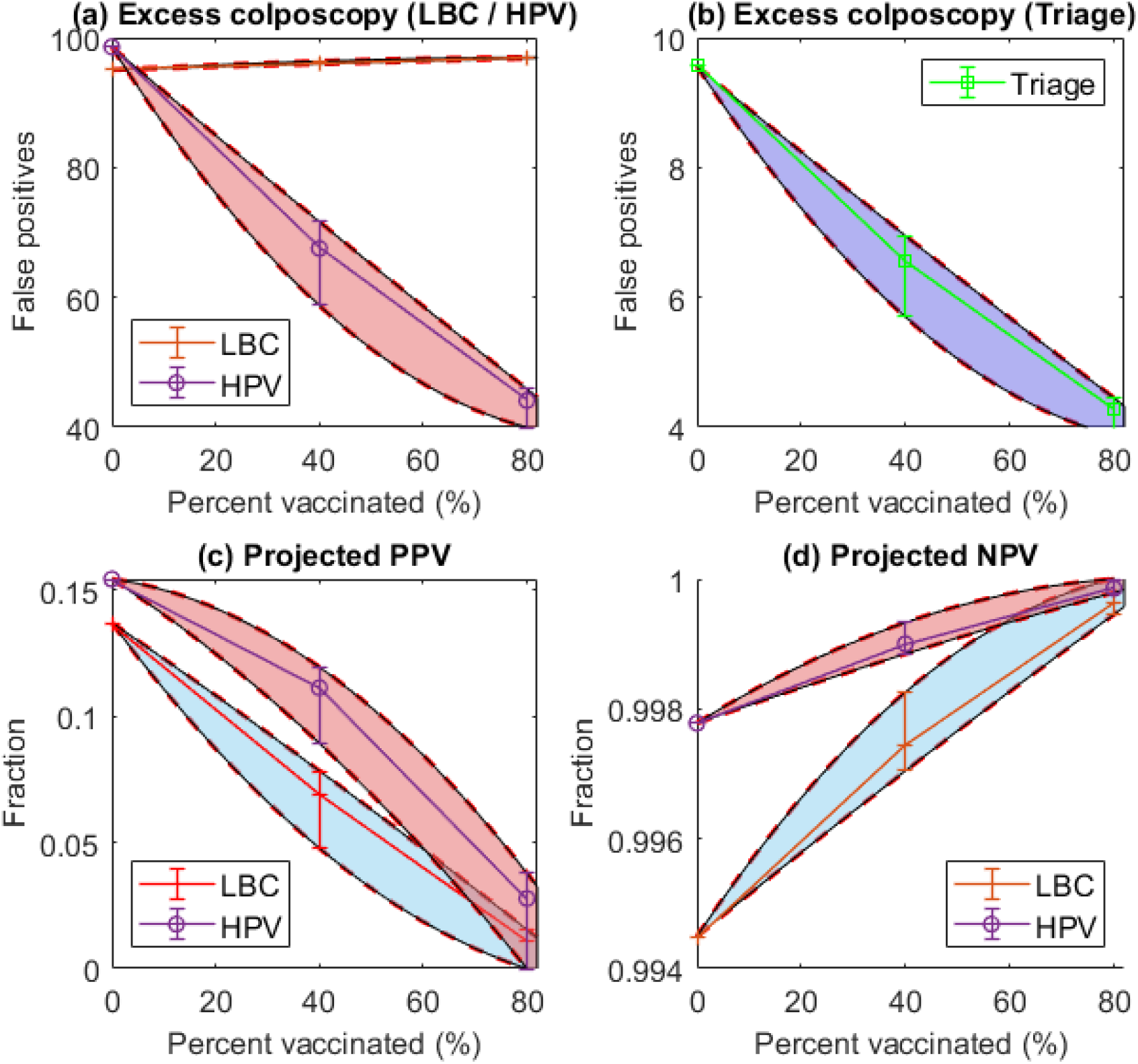
Impact of HPV vaccine uptake rates on (a) Number of excess colposcopy referrals (false positives) for both LBC and HPV screening (with caveat that HPV-only screening is not typically performed and is included for completeness) (b) Triage screening (c) Change in PPV with vaccination rate (d) Change in test NPV with vaccination rates. The envelopes in each figure refer to the range of estimates from 29 different models(16).

## Discussion

Cervical screening is a life-saving intervention, but as the results of this analysis show, it must be applied judiciously to have maximum benefit, whilst minimising impacts of over-treatment in false positive cases. In this work, we examined advantages and limitations of different screening methods. The improved confidence gained for the patient in a negative primary HPV result in comparison with cytology testing alone (performed at an equal frequency) is due to the HPV test’s improved sensitivity(27,28). The minimal benefit accrued in terms of sensitivity for treatable cervical cancers from co-testing, as outlined in this work, means that the decision of which modality to use is not only a scientific question, but one of appropriate allocation of limited public health resources rather than a scientific one which screening strategy should be pursued(29). Primary HPV testing at a 3 year interval has been demonstrated to have at a minimum equivalence with 5 yearly cotesting(30). Co-testing has other drawbacks too, as outlined in the result section and later in this discussion.

Primary testing(21,31) produces an abundance of false positives, and this can certainly be reduced with triage testing, as depicted in Figure 3. Both triage modalities (HPV with LBC reflex or LBC with HPV reflex) have only 10% the false positive rate of LBC, detecting 90% of the cases LBC would detect. As these approaches yield essentially the same result, then a screening programme could implement as their resources allowed without ill-effect. More importantly, triage outcomes are the same regardless of the primary test. As mentioned in the results section, this has economic implications. While the total number of tests required differs by only a small amount, if there is substantial differential costs between both modalities, then the optimum could be selected to minimise these without causing harm. If, for example, HPV tests were much costlier than LBC, then taking LBC primary approaches for triage would be cost-saving. Alternatively, if cytology was a limiting resource, then a HPV primary approach might be more suitable. A move towards HPV testing modalities will likely be strengthen given the WHO’s commitment to cervical cancer elimination.

The benefit of triage testing is the reduced number of excess colposcopies performed, but this comes at a slightly reduced detection rate. One potential approach to increase detection is to perform surveillance and expedited retesting of triage-negative women, and our results show that this approach should allow to eventually detect most prevalent CIN2/3 cases. Another potential approach to maximise detection rate is to perform co-testing, also shown in figure 2. This results in improved detection ratio relative to LBC primary (29% improvement) but at the cost of almost doubling the false positive ratio (95% increase). This is accordingly highly likely to prove excessively expensive, and ultimately detrimental to public health, as the increased rate of detection is associated with an amplified false positive rate. While this is a modality reduces missed cases of CIN2/3 cells, this analysis suggests it would not be viable, resulting in needless harm, as other authors have warned(32). This raises important ethical questions regarding the safety of any screening program. When an asymptomatic population are invited into a screening program, there remains an ethical obligation that they exit the program with a reduced cancer risk and minimal harm. Increasing referrals to colposcopy is likely to lead to over-treatment of dysplastic lesions with associated impact on fertility and obstetric outcomes, including a 2 fold increased risk of preterm birth(33). Over-diagnosis resulting from screening has long been recognised as a serious issue(17,34) with screening programmes, although it is one that remains difficult to quantify(35). The results of this work should be useful in elucidating potential harms and benefits.

While the model presented here is useful for quantifying detection statistics, it is important to consider the limitations of this analysis. For the false positive and negative life-time probability, we did not model natural history, and there is an implicit assumption that test results are independent from previous test results. The model is not adapted to predict the accuracy of screening at different ages, or for assessing the risk of progression/regression of CIN between screening opportunities. However, the results are likely a good approximation for the worst case scenarios, i.e. the risk for a woman with persistent CIN2/3 and the risks of screening for a woman who remains negative throughout her screening journey. This important assumption requires consideration as it is plausible that there are simply some CIN2/3 lesions that may never be detected with cytology or HPV testing due to characteristics of the lesion, such as low volume or low viral load. This influences the cumulative probability of a false negative/positive.

A crucial point to acknowledge is that all screening modalities have inherent limitations - those which maximise detection are most likely to lead to false positives, and those which reduce the incidences of false detection also reduce CIN2/3 detection. However, there are serious caveats to this that must be considered. Considering HPV triage with LBC screening in table 4, it is immediately apparent that expedited re-testing of HPV positive results outside of the regular screening cycle (6-18 months) goes a long way towards ameliorating the reduced detection ratio of triage tests, whilst minimising false positives and excess colposcopy referrals. This analysis also suggests that LBC only retesting of triage results tends to detect less disease than HPV retesting, or both HPV and LBC retesting.

In designing a screening programme, one must be cognisant of the potential harms as much as benefits. The advent of HPV testing has huge implications for cervical screening(11) (36), the implications of which are quantified further in this paper. The question of testing intervals was beyond the scope of this work, but was briefly alluded to in the analysis of false positive and false-negative cumulative probability illustrated for 1 year, 3 year, and 5 years intervals in figures 2 and 3. A recent French study(37) found that reducing testing window interval does more harm than good, leading to over-screening with needless risk, excess costs, and over-treatment. Other authors(28) have suggested that re-screening after a negative primary HPV screen should occur no sooner than every 3 years, and with Dillner et al^29^ reporting that intervals of even 6 year were safe and effective. 2018 US guidelines for HPV screening recommend a minimum interval of 5 years (26) between routine screening tests. As this analysis illustrates, we would expect in most cases that extending this interval has only a miniscule impact on missed CIN2/3 rate, while substantially reducing false positives.

As HPV testing becomes cheaper and more common, it is vital to consider how they are best implemented into screening. Evidence from recent multicentre studies(5,38) indicate that HPV-based screening provides greater protection against invasive cervical carcinomas relative to cytology. In those studies, recorded cumulative incidence of cervical cancer was lower 5.5 years after a negative HPV test than 3.5 years after a negative cytology result. This indicates empirically that 5-year intervals for HPV screening are safer than 3-year intervals for cytology. Results in this work support the hypothesis that HPV screening every 5 years could reduce the number of unnecessary colposcopy and biopsy procedures compared to frequent cytology, cutting costs and invasive unnecessary procedures. There is also ample evidence that negative hrHPV tests provide greater reassurance of low abnormality risk than negative cytology results(28,39), with authors suggesting that primary hrHPV screening can be considered as an alternative to current US cytology-based cervical cancer screening methods. Certainly, the results of this analysis support the contention that HPV testing can strongly increase predictive power of screening tests, and when correctly deployed can also reduce potential harms of over-screening.

It is vital to look towards the future of cervical screening. The staggering international success of the HPV vaccine is already apparent(40), and countries with high update of the HPV vaccine are already seeing a precipitous drop in rates of precancer and abnormal cervical cells. The falling prevalence of CIN2/3 has deep implications for how we interpret future tests. As figure 3 demonstrates, the chief impact of falling HPV infection rates is that across all modalities, positive results are less likely to be informative. Conversely, our confidence in negative results increases as vaccine rates increase. It is clear that HPV testing is superior as infection rates fall, and results in much fewer false positives relative to LBC testing. This is likely to be important in planning the future evolution of screening programmes. The model outlined in this work has application here too, and can be employed to predict the confidence one should afford a particular screening result under varying levels of population prevalence.

The requirement to both educate women and provide accurate information on the impact of any alteration to screening programmes can be illustrated by the psychosocial impact the addition to or replacement of LBC with HPV for primary screening or for triage which can cause additional stress and anxiety for those participating in the screening programme(41–43). Unfortunately the discrepancy between society’s expectation of screening programmes and actual sensitivities exist demonstrating the importance of public education(44). It is worth noting too that physicians and healthcare professionals are frequently under informed about the benefits and limitations of screening programmes(45,46), and confusion can easily arise. While screening is an extraordinary measure that saves lives, it is important to understand its fundamental limitations and nuances, so that maximum benefit can be derived from any national programme and misunderstandings minimised.

Cervical screening comes with some inherent uncertainty, irrespective of the modality employed. While this work should help elucidate some optimum strategies for screening, the reality is that screening, while life-saving, cannot be expected to be perfect. It is worth being clear that perfect detection is a mathematical impossibility, and this is demonstrated in the appendix. There is an inherent trade-off in strategies to increase detection, as they inevitably lead to a disproportionate rise in false positives, with needless over-treatment. This is particularly relevant in the context of the legal requirement in some jurisdictions, such as Ireland where following legal action over missed cancer diagnosis, the high court ruled that screeners must have ‘absolute confidence’ in negative results(10), despite multiple investigation showing the labs in question were operating to high standard and no negligence was committed.

Such a stipulation is impossible, and as this analysis shows, even striving to get arbitrarily close to this standard is likely to result in more harm than good. This is neither conducive to public health, nor sustainable. It also has potential to muddy public expectation and understanding of screening, and what it can realistically achieve. Screening is a vital undertaking if we are to reduce cervical cancer mortality, and its strengths and limitations must be seen in context so that benefit can be maximised. The results of this analysis should prove useful in optimising approaches and demonstrating the complexities of different implementations so informed decisions can be made.

### Electronic supplementary material

Model details, parameter estimation, and the equations used in this paper can be found in the electronic supplementary material.

## Data Availability

Supplied with paper

## Notes

### Competing Interest Statement

The authors have declared no competing interest.

### Funding Statement

No specific funding

